# Design, Implementation and Feasibility of an Informatics Infrastructure for Standardized Data Acquisition, Transfer, Storage and Export in Psychiatric Clinical Routine

**DOI:** 10.1101/2020.12.21.20248639

**Authors:** R. Blitz, M. Storck, B.T. Baune, M. Dugas, N. Opel

## Abstract

**Background:** Empirically driven personalized diagnostic and treatment is widely perceived as a major hallmark in psychiatry. However, databased personalized decision making requires standardized data acquisition and data access, which is currently absent in psychiatric clinical routine.

**Objective:** Here we describe the informatics infrastructure implemented at the psychiatric university hospital Münster allowing for standardized acquisition, transfer, storage and export of clinical data for future real-time predictive modelling in psychiatric routine.

**Methods:** We designed and implemented a technical architecture that includes an extension of the EHR via scalable standardized data collection, data transfer between EHR and research databases thus allowing to pool EHR and research data in a unified database and technical solutions for the visual presentation of collected data and analyses results in the EHR. The Single-source Metadata ARchitecture Transformation (SMA:T) was used as the software architecture. SMA:T is an extension of the EHR system and uses Module Driven Software Development to generate standardized applications and interfaces. The Operational Data Model (ODM) was used as the standard. Standardized data was entered on iPads via the Mobile Patient Survey (MoPat) and the web application Mopat@home, the standardized transmission, processing, display and export of data was realized via SMA:T.

**Results:** The technical feasibility was demonstrated in the course of the study. 19 standardized documentation forms with 241 items were created. In 317 patients, 6,451 instances were automatically transferred to the EHR system without errors. 96,323 instances were automatically transferred from the EHR system to the research database for further analyses.

**Conclusions:** With the present study, we present the successful implementation of the informatics infrastructure enabling standardized data acquisition, and data access for future real-time predictive modelling in clinical routine in psychiatry. The technical solution presented here might guide similar initiatives at other sites and thus help to pave the way towards future application of predictive models in psychiatric clinical routine.

## 1. Introduction

### 1.1 Scientific background

Psychiatric disorders represent one of the leading causes of disability worldwide. In the challenge to provide advanced treatment and prevention strategies for psychiatric disorders, previous research has focused on a better understanding of the neurobiological basis of affective disorders [1]. Yet the translation of such findings into clinical application remains an unresolved problem up to now. For this reason, the focus of psychiatric research has shifted from sole neurobiological characterization at the group-level towards the application of multivariate machine learning methods trained on multimodal data for an individualized prediction of clinical outcomes [2,3]. Multivariate machine learning (ML) applications have been proven innovative and powerful tools in translational psychiatric research. In this regard, the successful utilization of machine learning algorithms for individualized predictions of treatment response [4-6], depression severity [7], disease risk [8], differential diagnosis [9,10] and relapse risk [11] has yielded first promising results.

Yet, up to now, several obstacles prevent the successful transfer of individual predictive modeling to clinical routine application as discussed in recent reviews [12-15]. In this regard, the gap between homogeneous, well characterized samples acquired in experimental studies [16] and heterogeneous, unvalidated data from day-to-day clinical routine has proven to be a major obstacle in the translation of predictive models to clinical application. Hence, ecologically valid predictive models would require access to standardized real world data collected at the point-of-care [17].

Importantly, large-scale studies reporting the successful application of multivariate models trained on data from Electronic Health Record (EHR) including features such as diagnosis and procedures, laboratory parameters and medication for the prediction of suicide risk or weight gain following antidepressant treatment have demonstrated the capacity and generalizability of predictive models trained on real-world data [18-20]. Further extension of EHRs via standardized collection of predictive variables such as known risk factors might further enhance the potential of this novel data entity for predictive analytics in psychiatry [21,22]. Standardized electronic collection of patient-reported outcomes, that has previously been shown to improve clinical outcomes such as survival in cancer patients represents another possibility to enrich EHR data. Similarly, combining data from EHRs with research data might provide new opportunities for the discovery and validation of psychiatric endophenotypes as demonstrated via recent validation of a polygenic risk score in a danish population study [23].

However, future application of predictive models for personalized diagnostic and treatment requires their validation via clinical trials that in turn critically depend on the availability of the informatics infrastructure for an application of predictive models in routine care. The required informatics infrastructure should facilitate the acquisition of standardized real world data at the point-of-care, potential enrichment with patient reported outcomes or research data and subsequent access to data for clinicians and researchers. However, at present these technical requirements are widely absent in the clinical working environment.

The presented study aims to present the design and implementation of the technical requirements to address the aforementioned challenges with the ultimate goal of providing the basis for a successful future translation of predictive models to clinical application in psychiatric disorders. The implementation of the outlined technical solution shall ultimately allow to evaluate the potential of predictive models for the clinical management of psychiatric disorders under real-world conditions. In detail, we present the design and implementation of the informatics infrastructure including technical solutions for a) extension of EHR via standardized electronic collection of patient-reported outcomes, b) data transfer between EHR and research databases, c) pooling of EHR and research data in a unified database and d) visual presentation of analyses results in the EHR.

### 1.2 Objective of the study

The main objective of the presented study is the design and successful implementation of the informatics infrastructure required to train and validate predictive models in day-to-day clinical application in psychiatry as part of the SEED 11/19 study [24]. In detail, this includes the following steps

- Implementation of standardized documentation forms in electronic health records
- The set-up of an interface for direct data transfer between clinical documentation systems and a database for predictive analysis
- The set-up of a unified databases that allows pooling of clinical data with further research data for predictive analysis
- Visual presentation of relevant data entities and results of predictive analysis in electronic health records at the point-of-care

### 1.2 Setting

Münster University Hospital in Germany is a tertiary care hospital with 1,457 beds and 11,197 staff who treated 607,414 patients (inbound and outbound) in 2019 [25]. The department for psychiatry and psychotherapy at the University Hospital treated 1,341 cases in the study period from 25/02/2019 - 31/07/2020 (1,042 cases in 2018 [26,27]). Validation was carried out by 25 doctors and 61 specialists from the health care sector.

### 1.3 System details

The EHR system ORBIS by Dedalus Healthcare is used at Münster University Hospital in more than 40 clinics and is the market leader in Germany, Austria and Switzerland with over 1,300 installations [28]. The EHR system has an 8,700 GB Oracle database, 7,938 users and 1,927 user sessions per day (status at July 2020) at Münster University Hospital. No standardized metadata sets are supported.

### 1.4 Requirement engineering

To address the study aims outlined in the introduction, the following requirements were identified through focus groups including physicians and researchers at Münster University Hospital in Germany.

#### a) Extension of EHR via standardized data collection

At first sight the widely established usage of electronic documentation systems in clinical routine might supplement the notion of a fast translation of predictive models. Yet, up to date the majority of clinical data is still acquired and stored in an unstructured way that cannot be directly used for predictive modeling. Extension of EHR data via standardized forms of data collection in routine care is therefore required to provide a sufficient database for the development of predictive models. Importantly, the technical solution should be flexible and allow to update the content of collected EHR data.

Content-wise, in an initial step, standardized extension of EHR data should include assessment of symptomatology in order to allow both patient stratification at baseline as well as outcome measurement following intervention. Furthermore, standardized assessment of known risk factors including life events and sociodemographic data appears meaningful.

#### b) Data transfer

Routine EHR data storage systems are usually strictly separated from research databases for safety reasons and hence are not directly accessible for predictive analyses. Training and validation of predictive models based on EHR data requires the set-up of interfaces and a database in which EHR data can be transferred and subsequently stored in a standardized way. In line with our study aim, the technical solutions should be scalable and allow data transfer in real-time. EHR data transferred and stored in the database must be accessible for researchers in order to allow the development of predictive models.

#### c) Combination of EHR and research data

Again, since routine EHR data storage systems are strictly separated from research databases, pooling of EHR and research data is not possible within state-of-the art EHR databases. Pooling EHR with research data in a unified database would allow to enrich predictive models trained on EHR data by adding already existing research data and furthermore to validate EHR data based on research data. To this end, in order to combine each patients’ EHR and research data, a unified scalable research database is needed that allows to integrate EHR and research data acquired via experimental studies.

#### d) Presentation of standardized data within the EHR

Once collected, clinically useful standardized data as well as results of any analysis must be transferred back to the main EHR system in real-time and presented to the clinician at the point of care.

### 1.5 Solution requirements

An informatics infrastructure enabling real-time clinical predictive modeling based on the single-source architecture was derived from the named requirements. Custom metadata must be supported. Clinical Data Interchange Standards Consortium (CDISC) Operational Data Model (ODM) (version 1.3.2) was used as a flexible standard for exchange and archiving of metadata within the framework of clinical studies [29,30]. Mobile applications must be able to communicate with the architecture. Automatic data transfer into the database of the EHR system and from the EHR system to a research database was carried out via a communication server. ODM files were transported automatically to the database of the EHR system with Health Level 7 (HL7) messages [31]. NextGen© Connect [32] was used as a communication server. HL7 version 2.5 and message type ORU^R01 were used. Plausibility and completeness of form data had been validated by clinical users.

## 2. Methods

### 2.1 Analysis of technical and clinical feasibility

The technical feasibility was demonstrated by the implementation of an infrastructure that enables clinical predictive modeling in real time. Java version 1.8.0_181 [33], JavaScript ECMAScript 6 [34], TypeScript version 3.7.2 [35] and the proprietary language of the EHR system were used as programming languages. MongoDB Java Drivers version 3.9.1 [36] and Json-lib version 2.4 [37] were used as third party libraries. MongoDB version 4.2.3 [38] was used as a research database, Docker version 19.03.13 [39] for containering and Red Hat Enterprise version 7.8 [40] as research server. The clinical feasibility was determined by piloting the architecture in the clinic for psychiatry and psychotherapy and a prospective analysis of the clinical documentation forms used. 25 doctors and 61 health care sector specialists were clinical users of the system. Stakeholder of the study at Münster University Hospital is the Institute for Translational Psychiatry, Department of Psychiatry. Evaluation began on February 25, 2019 and ended on July 31, 2020. The following evaluation criteria were analyzed:

1. Measurement of data completeness in created documentation forms
2. Measurement of data completeness in the research database
3. Monitoring of system stability
4. Monitoring of data transfer

IBM SPSS Statistics version 25 [41] was used for descriptive data analysis. Adobe Photoshop version 11.0 [42] and Microsoft Visio version 16.0.4849.1000 [43] were used to depict the workflow.

## 3. Results

### 3.1 System architecture

The single-source metadata architecture transformation (SMA:T) was used as the software architecture [44]. SMA:T is an extension of the EHR system of the Münster University Hospital and uses Module Driven Software Development [45] to generate standardized applications and interfaces. Every SMA:T form has a generic built-in interface for exchanging standardized data. Embedded applications [44] were used as the application type. These are linked to an Operational Data Model (ODM) file in the EHR database, from which a documentation form is generated. All metadata and clinical data are available in the Operational Data Model (ODM) developed by CDISC in version 1.3.2. Patient reported outcomes are recorded via Mobile Patient Survey (MoPat) [46,47] on mobile devices (generation 6 iPads) and via the web application Mopat@home (a modified version of the tablet-based web application MoPat) [48] for follow-up assessments following discharge from inpatient treatment. Collected data are transferred to the communication server via an HL7 message and from there to the database of the EHR system. Data are sent in the OBX-5 segment of the HL7 message. SMA:T provides database storage. A reference to the imported clinical data is saved. Clinical data are automatically inserted by SMA:T when the documentation form is opened for the first time. A unique ID from the HL7 header is used for this purpose. Each ID is linked to an imported clinical record. The structure of the architecture is shown in Figure 1. Data transfer to the research database takes place via the researcher module from SMA:T. This provides a front end to the EHR system and an extension of the communication server for data transfers. Both prospective and retrospective standardized data exports of EHR data points are supported, specifically vital signs, laboratory data, medication data and administrative data. Each data export can be customized by individual parameters. The following parameters are supported: Name of data export, export interval, database query, destination parameters for Electronic Data Capture (EDC) systems or research databases. MongoDB and RedCap [49] are currently provided as destination templates in the EHR system. The destination portfolio can easily be expanded with interface functions of SMA:T. The research database is embedded in a Docker container of a virtualized Red Hat Enterprise Linux server. The data flow from EHR to EDC is shown in Figure 2. The software architecture is shown in Figure 3.

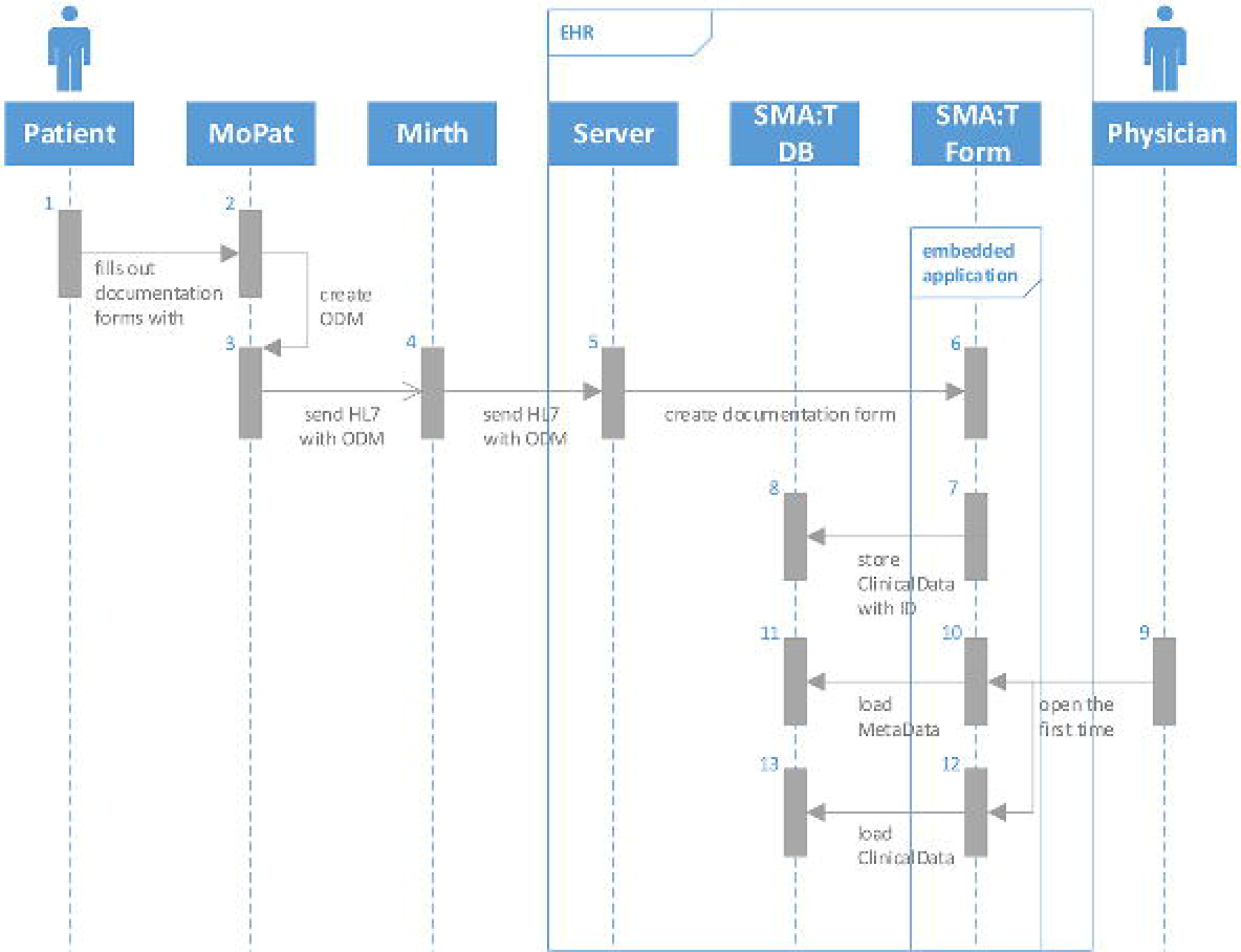

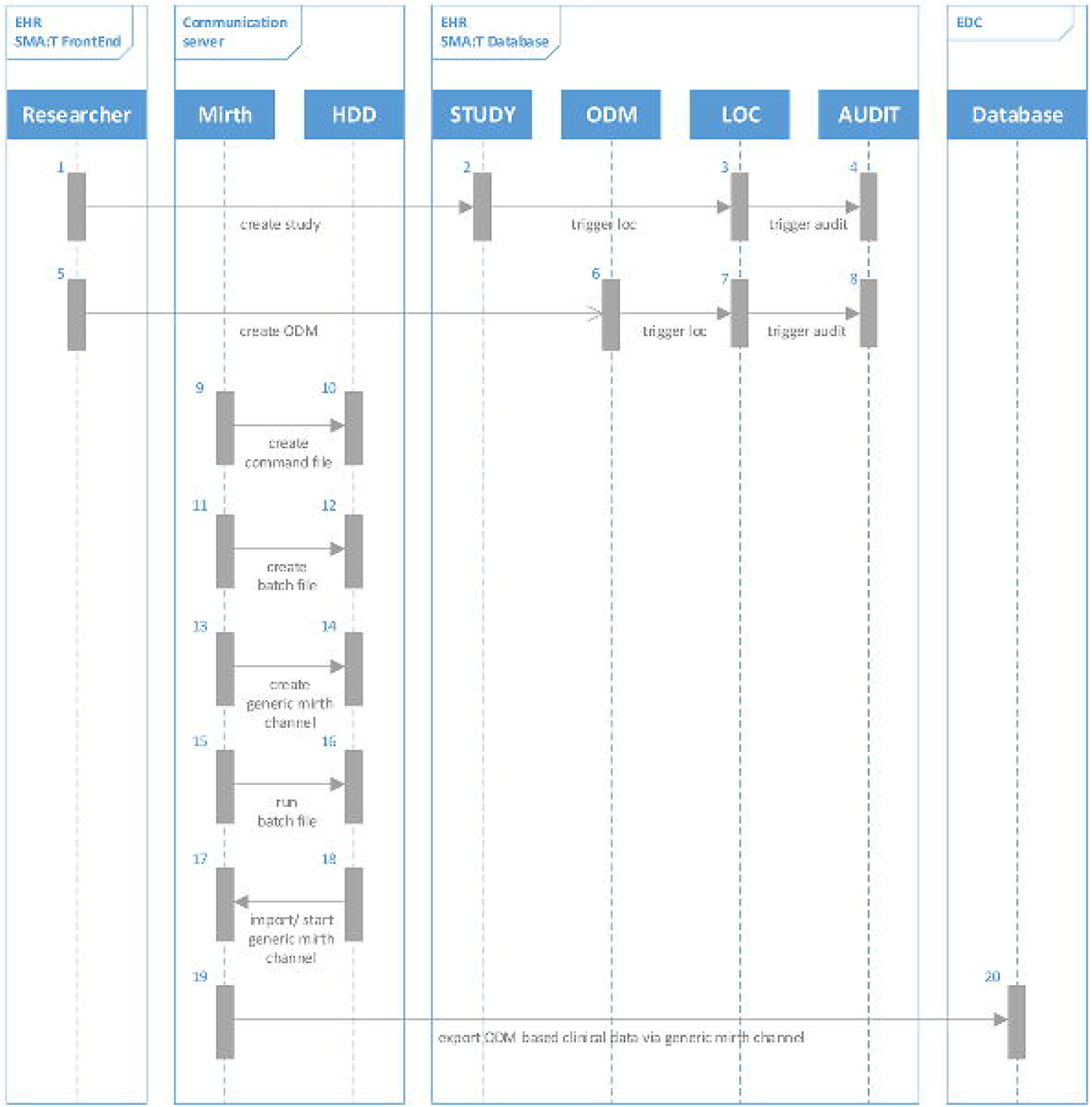

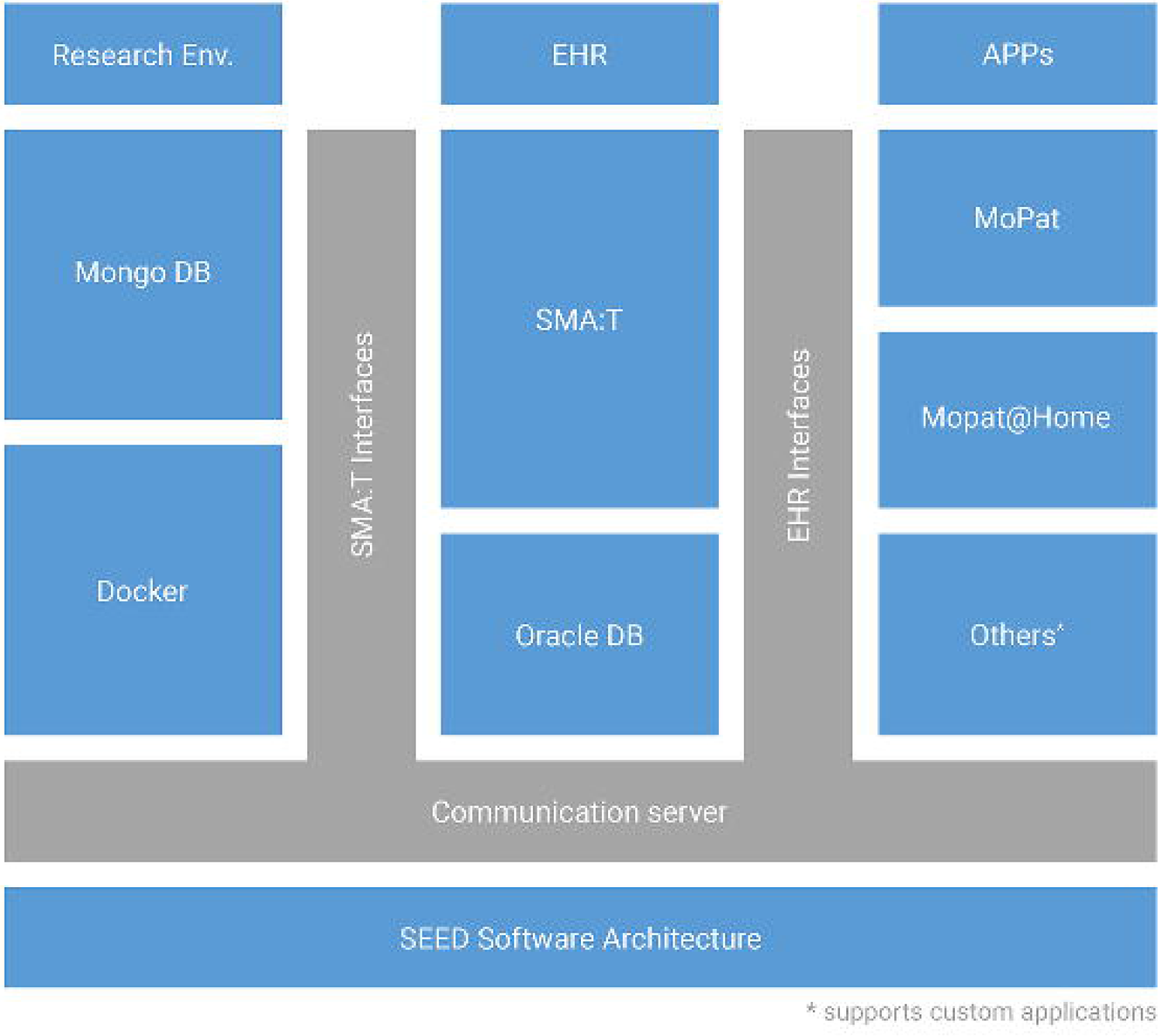

### 3.2 System implementation

The implementation of the architecture is divided into four areas: data collection, data transfer, data storage and data visualization. Agile methods were used for Project Life Cycle and Development Cycle [50].

#### a) Data collection

SMA:T provides two options for data collection: EHR system in clinical routine and dedicated web applications. Data input via web applications can be designed freely. In the present study, EHR data generated as part of clinical routine documentation comprised among others laboratory data, medication, information on diagnosis, time of admission, length of stay and are presented in detail in Table 1. MoPat [46,47] was selected for the collection of patient-reported outcomes. After input of patient case ID, staff handed the patient an iPad with the MoPat app. Patients were then guided through a series of documentation forms comprising different questionnaires and entered data on the mobile device (Details in Figure 4). The iPad was then returned to the medical staff. Further details regarding the collection of patient-reported outcomes during inpatient treatment have previously been described [24]. In addition, Mopat@home was used for the collection of patient-reported outcomes following discharge. To this end, patients were send an e-mail, which provided a link to a website on which the above referenced questionnaires were presented and could be filled out [48].

**Table 1.** Research documentation used in the Department of Psychiatry. 8 Documentation forms with 39 Items were developed for the standardized retrospective export of the study.

| Name | #Items |
| --- | --- |
| SEED ClinicalData Admission Date & Time | 4 |
| SEED ClinicalData Classification | 2 |
| SEED ClinicalData DRG / Diagnosis | 3 |
| SEED ClinicalData ECT | 11 |
| SEED ClinicalData Laboratory Assessments | 7 |
| SEED ClinicalData Medication | 5 |
| SEED ClinicalData Patient | 4 |
| SEED ClinicalData Vital Signs | 3 |

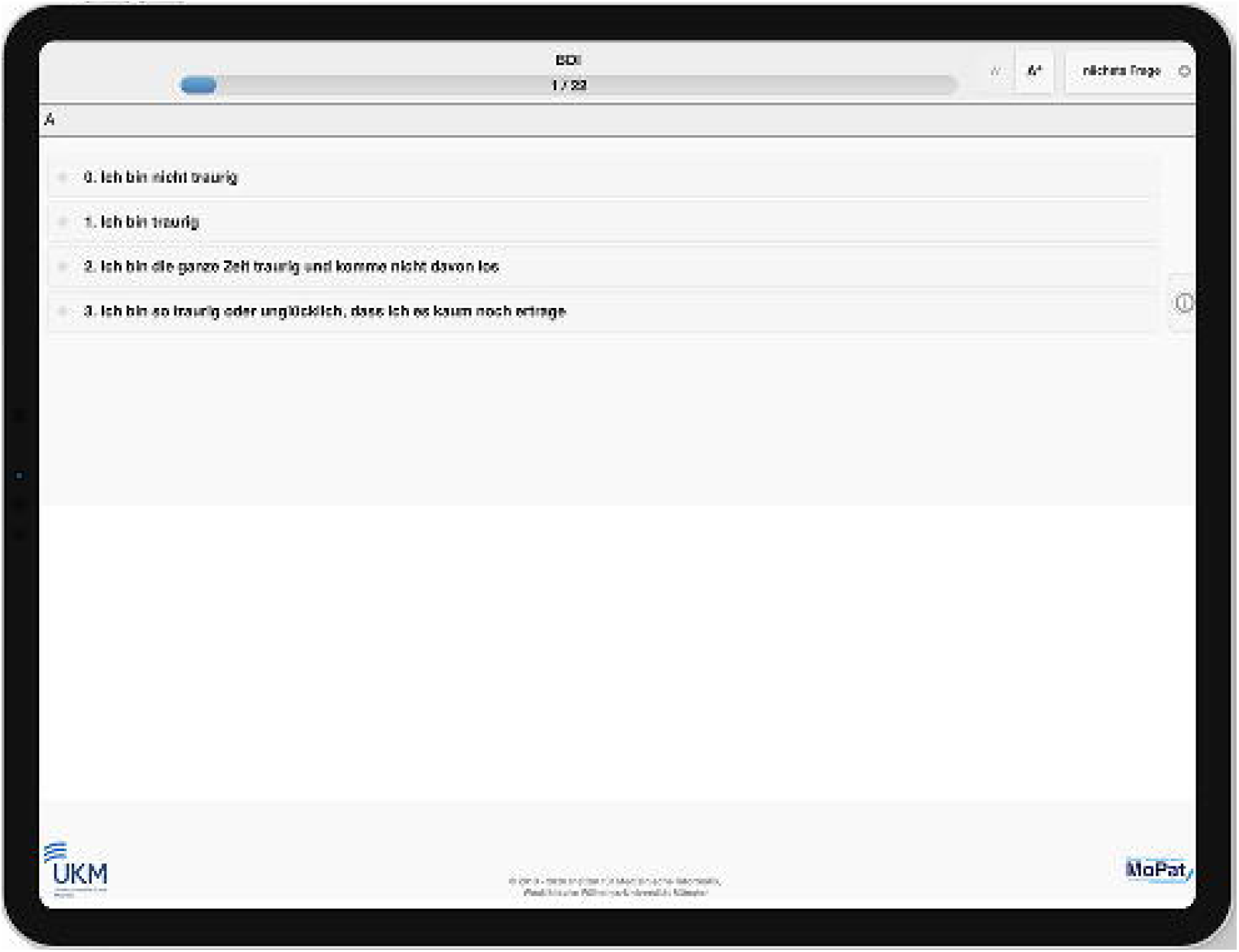

#### b) Data transfer

SMA:T provides two types of data transfer in the present scenario: data transfer into the EHR system and transfer into the EDC system. MoPat sends data to the EHR system via the communication server of the University Hospital. Data are saved in the ClinicalData structure of the ODM format. The ODM document is embedded in an HL7 message. Each HL7 message creates a form in the EHR system. The header of the HL7 message determines which form is automatically created. Data transfer to the EDC takes place via SMA:T interfaces. Both retrospective and prospective data exports in real time are supported. When a study query was activated via the EHR frontend, metadata and corresponding SQL statement are read by the SMA:T extension of the communication server. SMA:T uses its code library and channel framework to generate unique Mirth channels. These send a database query to the EHR system and transfers the output directly to the EDC system. Both metadata (clinical documentation form) and clinical patient data are provided by SMA:T in ODM format. Data records are combined into an ODM document. In the present study, SMA:T converts the resulting XML-based ODM document into JSON format [51] (JODM format [52]). The JSON schema [53] of JODM is open source and currently limited to Study and ClinicalData nodes, including all sub-nodes of the Operational Data Model in version 1.3.2.

#### c) Data storage

Data storage addresses metadata and clinical data. Metadata of clinical documentation forms are stored centrally in the SMA:T database. The SMA:T database model is part of the EHR database model. Metadata and clinical data are available in ODM format. MoPat also supports ODM format; thereby the same data model can be used for both systems. Clinical data are clearly identified by unique OIDs and the associated OID on the documentation form.

#### d) Data presentation

Usability principles were applied to visualize data [54-56]. A 1-column layout was implemented according to the requirements of the ten Web Form Design Guidelines [57].Those forms are displayed via SMA:T within the EHR system (see Figure 5, Figure 6). SMA:T supports item-based real-time notifications as well as centralized notification services to display analysis results in real time.

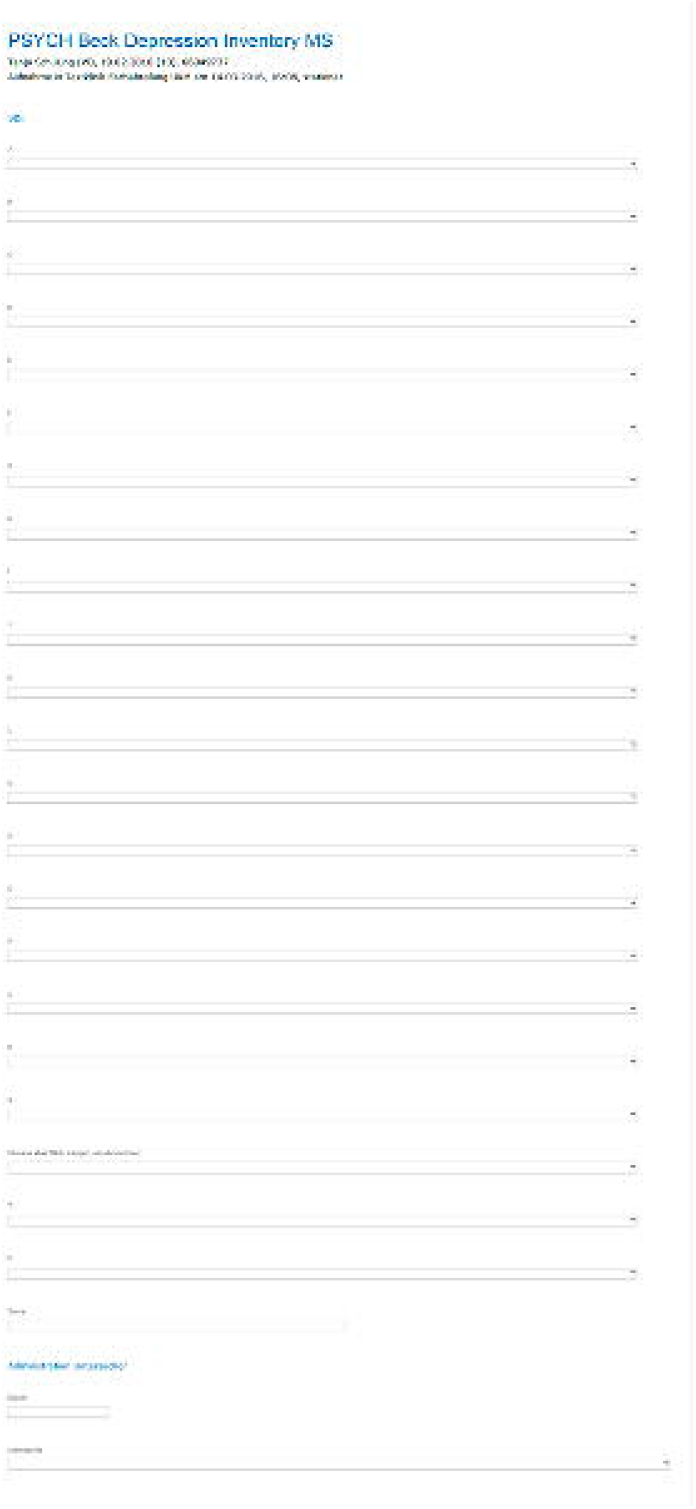

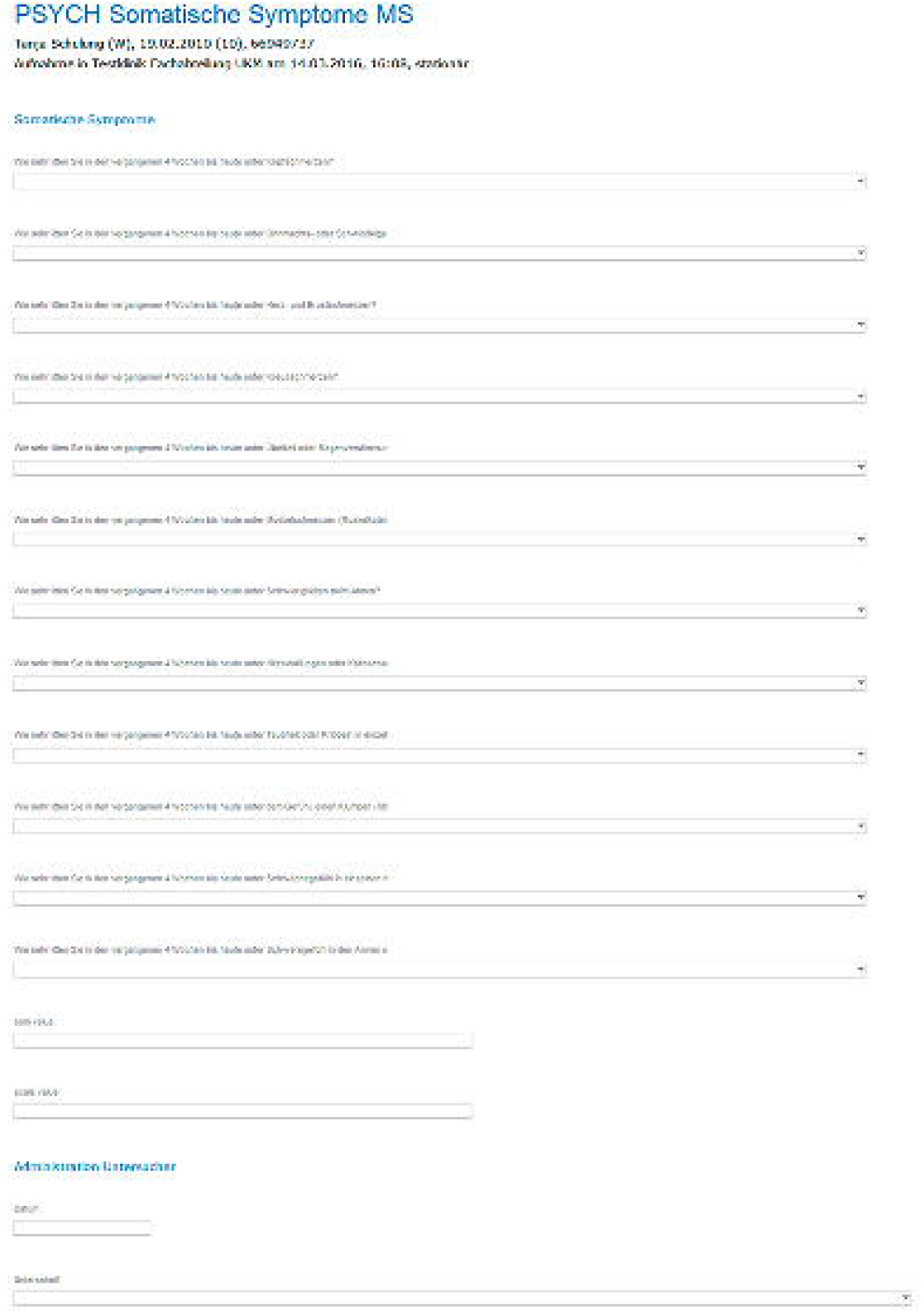

### 3.3. Technical and clinical feasibility

As part of the study, 11 standardized documentation forms with 202 items were created for the clinic for psychiatry and psychotherapy (Table 2). 5,866 instances were created by the patients and automatically transferred to the EHR system of the University Hospital Münster without errors. 412 cases from 317 patients were processed by 86 users (Table 3). 86/123 (70%) of the medical staff of the clinic for psychiatry and psychotherapy worked with those documentation forms. Automatic generation of documentation forms was accepted in routine clinical use. Standardized data transfer from the communication server into the EHR system was completed without error. It was possible to display all items (n=202) from ODM structures in full using the generic workflow. Clinical data from 317 patients was stored in the EHR database. 96,7% of 4,509 scores could be calculated and transferred into the EHR system (Table 4). 111,842 items were completed by patients on mobile devices (Table 5). 99.3% of forms with scores (#Items 91,974 / #uncompleted items 645) were completed; 88% of forms without scores (#items 14,318 / #uncompleted items 1,701). Validity of the aquired data on depressive symptomatology was already analyzed in a feasibility study by Richter et al. [24]. 8 standardized documentation forms with 39 items were created for the retrospective data export (Table 1). A total of 96,323 instances of vital signs, laboratory data, medication data and administrative data could be automatically transferred from the EHR system to the research database (Table 6). Retrospective ODM-based data export worked correctly without technical errors. 585 instances were created by the patients with Mopat@home and transferred to the research database via SMA:T (Table 7).

**Table 2.** Routine documentation used in the Department of Psychiatry. 11 Documentation forms with 202 Items were developed for the study. Data models without license restrictions are available in the portal of Medical Data Models (MDM).

| Name | #Items | URL |
| --- | --- | --- |
| Beck Depression Inventory | 23 | https://medical-data-models.org/27300 |
| Big Five Inventory (BFI-2-S) | 35 | https://medical-data-models.org/41674 |
| Big Five Inventory (BFI-2-XS) | 20 | https://medical-data-models.org/41673 |
| Childhood Trauma Questionnaire (CTQ) | 34 | https://medical-data-models.org/9946 |
| Family Mental History | 14 | https://medical-data-models.org/41444 |
| Hamilton Depression Scale | 25 |  |
| NARQ-S | 9 | https://medical-data-models.org/41440 |
| SCL-90 Somatisation Scale | 14 | https://medical-data-models.org/41671 |
| Sociodemographic Questionnaire | 5 | https://medical-data-models.org/41441 |
| Questions on individual disease course | 18 | https://medical-data-models.org/41442 |
| Questions on somatic comorbidities | 5 | https://medical-data-models.org/41443 |

**Table 3.**
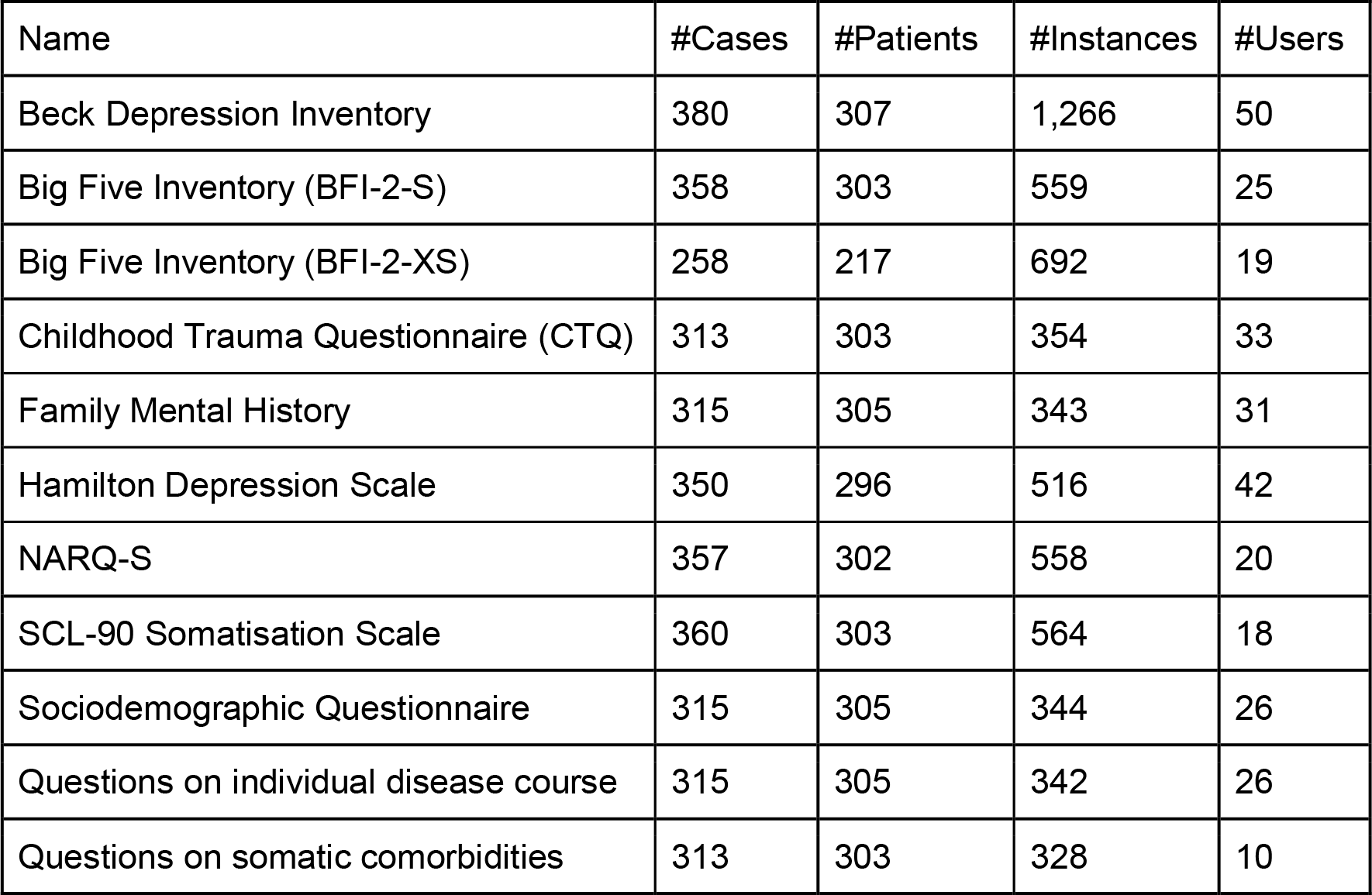
Number of instances created for each documentation form. Counts of patients, patient cases and users are shown.

| Name | #Cases | #Patients | #Instances | #Users |
| --- | --- | --- | --- | --- |
| Beck Depression Inventory | 380 | 307 | 1,266 | 50 |
| Big Five Inventory (BFI-2-S) | 358 | 303 | 559 | 25 |
| Big Five Inventory (BFI-2-XS) | 258 | 217 | 692 | 19 |
| Childhood Trauma Questionnaire (CTQ) | 313 | 303 | 354 | 33 |
| Family Mental History | 315 | 305 | 343 | 31 |
| Hamilton Depression Scale | 350 | 296 | 516 | 42 |
| NARQ-S | 357 | 302 | 558 | 20 |
| SCL-90 Somatisation Scale | 360 | 303 | 564 | 18 |
| Sociodemographic Questionnaire | 315 | 305 | 344 | 26 |
| Questions on individual disease course | 315 | 305 | 342 | 26 |
| Questions on somatic comorbidities | 313 | 303 | 328 | 10 |

**Table 4.**
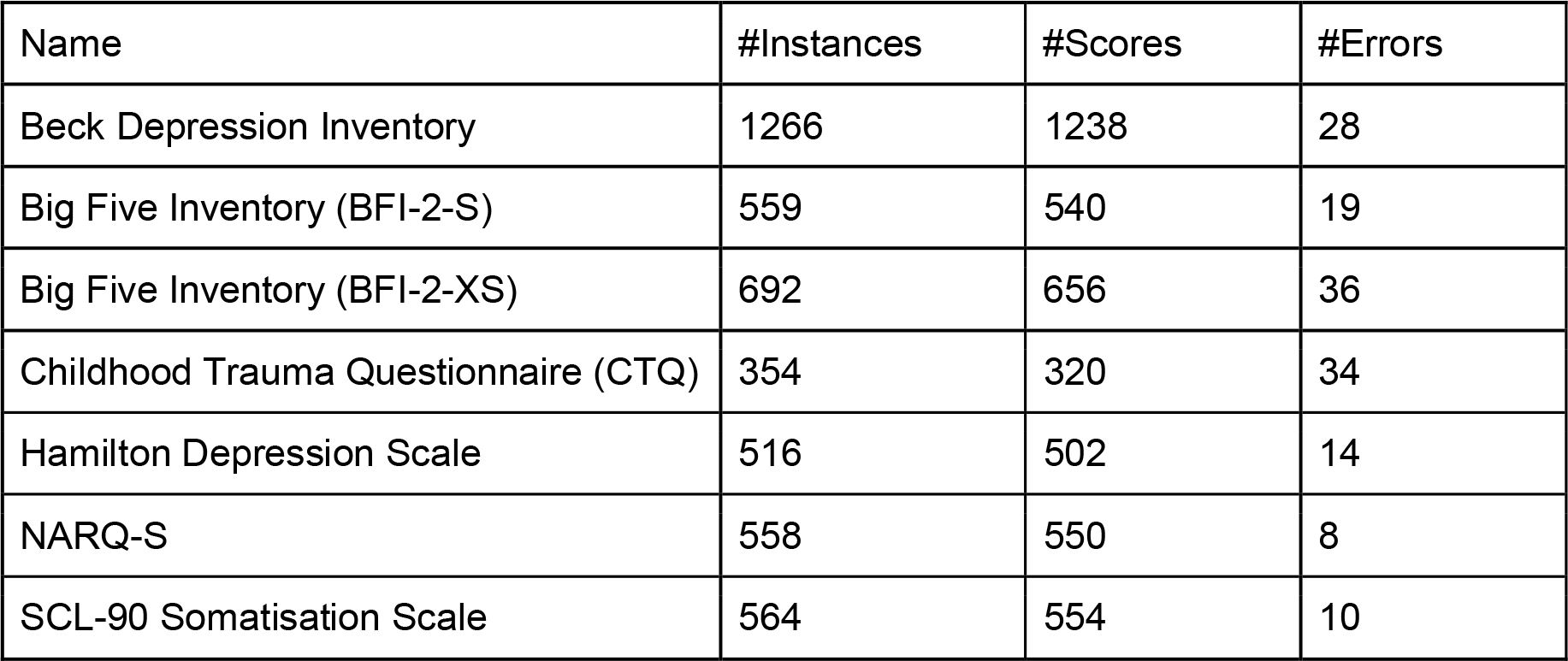
Data quality of patient based documentation regarding score calculation. Error frequency is determined by missing data entries.

| Name | #Instances | #Scores | #Errors |
| --- | --- | --- | --- |
| Beck Depression Inventory | 1266 | 1238 | 28 |
| Big Five Inventory (BFI-2-S) | 559 | 540 | 19 |
| Big Five Inventory (BFI-2-XS) | 692 | 656 | 36 |
| Childhood Trauma Questionnaire (CTQ) | 354 | 320 | 34 |
| Hamilton Depression Scale | 516 | 502 | 14 |
| NARQ-S | 558 | 550 | 8 |
| SCL-90 Somatisation Scale | 564 | 554 | 10 |

**Table 5.**
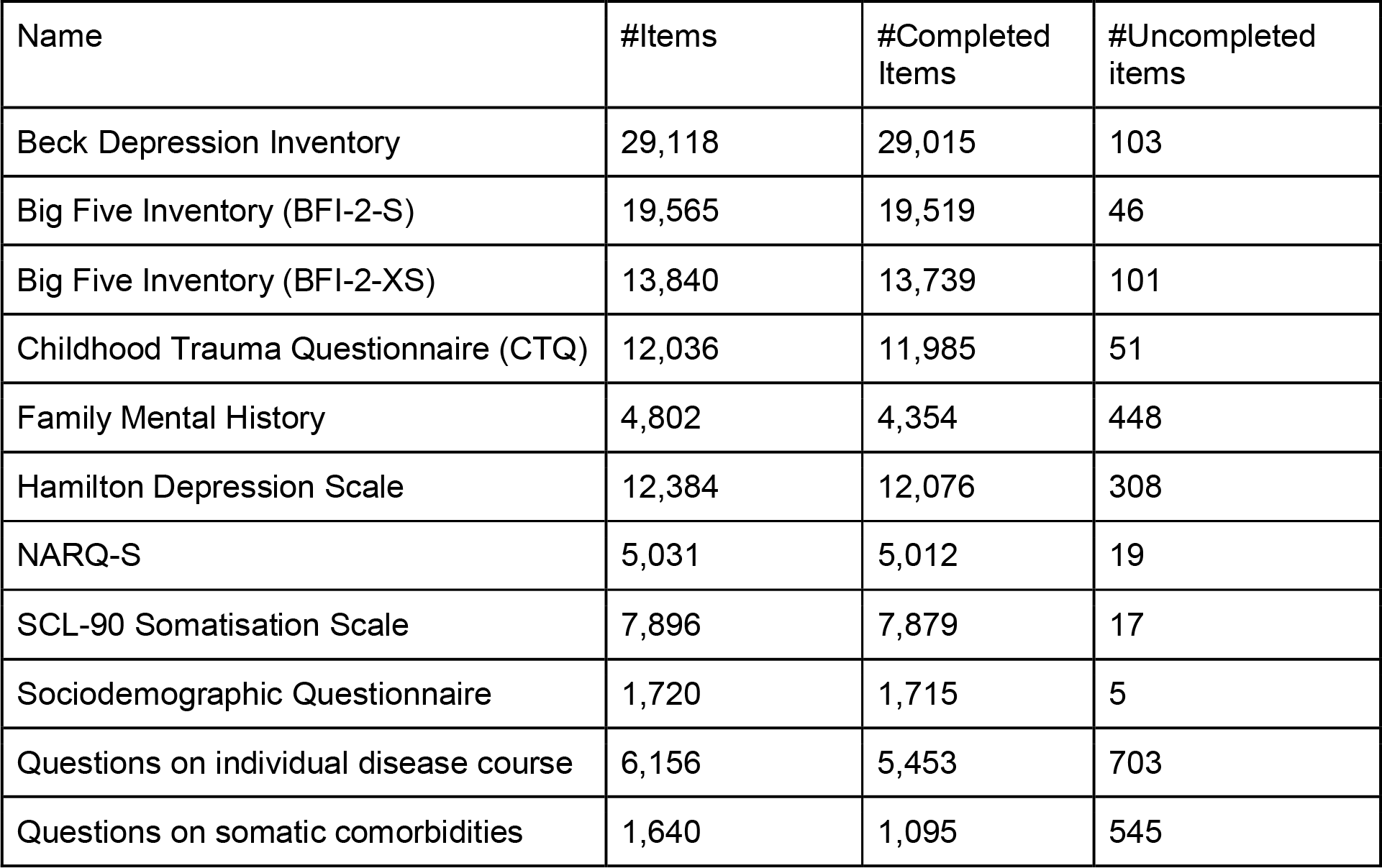
Completeness of documentation forms.

**Table 6.** Number of retrospectively transferred research documentation forms (EHR => EDC). EHR data was extracted with generic study queries in the SMA:T system.

| Name | #Instances (EHR) |
| --- | --- |
| SEED ClinicalData Admission Date & Time | 245 |
| SEED ClinicalData Classification | 8,260 |
| SEED ClinicalData DRG / Diagnosis | 1,163 |
| SEED ClinicalData ECT | 452 |
| SEED ClinicalData Laboratory Assessments | 22,886 |
| SEED ClinicalData Medication | 14,244 |
| SEED ClinicalData Patient | 245 |
| SEED ClinicalData Vital Signs | 48,828 |

**Table 7.** Number of instances created with Mopat@home for each documentation form.

| Name | #Instances |
| --- | --- |
| Beck Depression Inventory | 65 |
| Big Five Inventory (BFI-2-S) | 64 |
| Childhood Trauma Questionnaire (CTQ) | 65 |
| Family Mental History | 65 |
| NARQ-S | 64 |
| SCL-90 Somatisation Scale | 66 |
| Sociodemographic Questionnaire | 66 |
| Questions on individual disease course | 65 |
| Questions on somatic comorbidities | 65 |

## 4. Discussion

### 4.1 Answers to the study questions

The aim of the study was the design and implementation of the informatics infrastructure enabling standardized data acquisition at the point-of-care and subsequent accessibility of clinical data for analytic purposes which is required for future application of predictive models in day-to-day clinical routine in psychiatry. The overall technical feasibility of the implemented solution was shown. Standardized documentation forms were implemented to extend EHR data domains and to improve data quality in the EHR system. An automated transfer of data into the EHR and the research database was implemented thus allowing to pool EHR data with already existing research data from ongoing cohort studies. The system was accepted by clinical staff from the Department of Psychiatry. Widespread use of documentation forms could be demonstrated. Standardized electronic data collection in the EHR at the Point-of-Care was successfully implemented. The latter solution can similarly be applied for the presentation of results from predictive models.

### 4.2 Strengths and weaknesses of the study

Major strengths of this study include standardized acquisition, transfer, storage and export of data in real-time with a generic informatics infrastructure. This system fulfills prerequisites for future predictive modelling in clinical routine in psychiatry [58-60]. Standardized data transfer with ODM format provides scalability in the context of complex medical data structures. The Define-XML standard, an extension of the ODM standard, is mandated by regulatory authorities such as Food and Drug Administration (FDA) for metadata [61]. Compliance with regulatory standards is a major advantage of our infrastructure regarding future clinical studies. The data format had to be converted due to the research database, which is a limitation. MongoDB was chosen for real-time analysis of large amounts of data. Standardized automatic data transfer into research databases was possible for both retrospective and prospective research questions. The data of the EHR system was responsible for the number of documentation forms for the retrospective export. Data export can be configured centrally from the EHR system in compliance with local data protection regulations. Our approach is scalable, because ORBIS EHR systems are used in more than 1,300 hospitals in Germany, Austria and Switzerland. The evaluation concentrated on technical and clinical feasibility. Limitations include the lack of elaborated standardized evaluations of the user experience of the system by clinical staff. Moreover, further evaluation is necessary in order to assess the sustainable benefit in everyday clinical practice.

### 4.3 Results in relation to other studies

With the present study, we extend a previous line of research on predictive modeling based on EHR data. While previous studies have demonstrated empirical evidence for the predictive validity of EHR data in psychiatric use cases [18-20], to the best of our knowledge the present study is the first to not only report on the design but also on successful implementation and technical feasibility of the informatics infrastructure for standardized acquisition, transfer, storage and access of real world data for analytic purposes in psychiatric care which is the basic requirement for the application and validation of predictive models in future clinical studies.

While we are not aware of any other study that reported successful implementation of a comparable informatics infrastructure in psychiatric clinical routine, several preliminary reports should be taken into account. Complementary to the work presented in the present study, Khalilia et. al. described a Fast Healthcare Interoperability Resources (FHIR) web modeling service that was tested on a pilot ICU dataset [62]. A multi-source approach was used. No binding standard is used for clinical studies, instead the standard OMOP Common Data Model was applied [63] and a FHIR server and database are required for this system, which might limit potential implementations at multiple sites considering that many EHR systems currently do not yet use a FHIR server. Of note, we are aware of several large-scale efforts aiming to translate predictive models into psychiatric practice [64] that - once implemented - might serve as a future base for comparison of system stability and performance.

### 4.4 Generalizability of the study

The informatics infrastructure for standardized data acquisition, transfer, storage and export in real-time for future predictive modelling outlined in the present study is an important step in the complex process towards an implementation of machine learning and clinical decision support solutions in routine care. Our study shows that this approach is technically feasible. Due to the standardization, the concept is also scalable for other medical areas. Data warehouse applications of a heterogeneous hospital landscape can be implemented with this software architecture. In addition to local artificial intelligence applications, multi-site implementations of the architecture could also transfer pseudonymized data points into a global predictive model. The implementation of national and international predictive models in medicine would be possible.

### 4.5 Future work

Artificial intelligence systems rely on high quality data. In the future, AI applications might send real-time evaluations directly back into the EHR system. Clinical staff could access and respond to calculated predictions. Medical device regulation needs to be taken into account for implementation of such systems. Direct data transfer back from the clinic would be possible. Real-time adjustments of the prediction models would thus be possible. Standardization of clinical routine documentation via SMA:T can provide high quality structured data points. Specific prediction models can be trained in this way with the same architecture. Generic model pipelines can be set up. Model clusters can be set up to answer complex medical questions. Basically, SMA:T forms a solid technical infrastructure for the implementation of AI solutions in medicine. Scheme extensions of the ODM standard can be implemented to optimize communication between systems. Observational and interventional studies are warranted to evaluate predictive validity of machine learning models in psychiatric routine. For multi-center studies, SMA:T needs to be reimplemented in the respective EHR environments. A software blueprint is available [44]. Another important consideration is the potential future enrichment of EHR data with mobile assessments including ecological momentary assessments (EMA) and passive sensor data derived from smartphones. Recent reports on successful real-time prediction of depressive symptoms based on EMA data supplement this notion [65]. Thus, future studies should explore technical solutions that allow data transfer between EHRs and patients’ smartphone.

### 4.5. Conclusions

The presented informatics infrastructure enabling standardized data acquisition, transfer, storage and export in real-time for future predictive modelling in clinical routine in psychiatry is technically feasible. The outlined architecture provides a technical basis for the application, first and foremost the validation of clinical decision support systems and artificial intelligence applications in clinical studies.

## Data Availability

Authors can confirm that all relevant data are included in the article and/or its supplementary information files.

## 6. Abbreviations

CDISC: Clinical Data Interchange Standards Consortium
EDC: Electronic Data Capture
EHR: Electronic Health Record
EMA: European Medicines Agency
FDA: Food and Drug Administration
FHIR: Fast Healthcare Interoperability Resources H
DD: Hard Disc Drive
HL7: Health Level 7
JSON: JavaScript Object Notation
LOC: Lines Of Code
MoPat: Mobile Patient Survey
ODM: Operational Data Model
OID: Object Identifier
OMOP: Observational Medical Outcomes Partnership
SMA:T: Single-source Metadata ARchitecture Transformation
XML: Extensible Markup Language

## 8. Acknowledgements

We are deeply indebted to all participants of this study. Supported by a grant from BMBF (HiGHmed 01ZZ1802V).

